# Prolonged activation of nasal immune cell populations and development of tissue-resident SARS-CoV-2 specific CD8 T cell responses following COVID-19

**DOI:** 10.1101/2021.04.19.21255727

**Authors:** Anna H.E. Roukens, Marion König, Tim Dalebout, Tamar Tak, Shohreh Azimi, Yvonne Kruize, Cilia R. Pothast, Renate S. Hagedoorn, Sesmu M. Arbous, Jaimie L. H. Zhang, Maaike Verheij, Corine Prins, Anne M. van der Does, Pieter S. Hiemstra, Jutte J.C. de Vries, Jacqueline J. Janse, Meta Roestenberg, Sebenzile K. Myeni, Marjolein Kikkert, Mirjam H.M. Heemskerk, Maria Yazdanbakhsh, Hermelijn H. Smits, Simon P. Jochems, BEAT-COVID group

**Author notes:** Joint senior authors (Hermelijn H. Smits, Simon P. Jochems).

## Abstract

The immune system plays a major role in Coronavirus Disease 2019 (COVID-19) pathogenesis, viral clearance and protection against re-infection. Immune cell dynamics during COVID-19 have been extensively documented in peripheral blood, but remain elusive in the respiratory tract. We performed minimally-invasive nasal curettage and mass cytometry to characterize nasal immune cells of COVID-19 patients during and 5-6 weeks after hospitalization. Contrary to observations in blood, no general T cell depletion at the nasal mucosa could be detected. Instead, we observed increased numbers of nasal granulocytes, monocytes, CD11c^+^ NK cells and exhausted CD4^+^ T effector memory cells during acute COVID-19 compared to age-matched healthy controls. These pro-inflammatory responses were found associated with viral load, while neutrophils also negatively correlated with oxygen saturation levels. Cell numbers mostly normalized following convalescence, except for persisting CD127^+^ granulocytes and activated T cells, including CD38^+^ CD8^+^ tissue-resident memory T cells. Moreover, we identified SARS-CoV-2 specific CD8^+^ T cells in the nasal mucosa in convalescent patients. Thus, COVID-19 has both transient and long-term effects on the immune system in the upper airway.

## Main

Individuals infected with SARS-CoV-2 suffer from a wide range of symptoms, ranging from none to fever, cough and dyspnea and severe acute respiratory distress syndrome, which can culminate in death ^1^. The immune cell perturbations during COVID-19 have been described extensively in blood, with changes observed in almost all immune cell populations that often could be linked to disease severity. A global depletion of peripheral blood T cells has been proposed to be a hallmark of COVID-19 ^2,3^. At the same time, neutrophil numbers are strongly increased and an increased neutrophil to lymphocyte ratio is associated with poor prognosis ^4^. Natural killer (NK) cells show an activated profile, associating with disease severity ^5^. Myeloid cells show an aberrant profile with increased proliferation and altered functionality ^6,7^. The B cell compartment is characterized by oligoclonal expansion of plasmablasts and extrafollicular B cells ^8,9^. Eosinophils were found to be decreased in blood in most patients at time of hospital admission ^10^. However, eosinophils might expand during hospitalization and show upregulated levels of the homing marker CD62L and activation profiles, which was found to precede lung hyperinflammation ^11^.

Although SARS-CoV-2 mainly replicates in the respiratory tract, and lower respiratory tract complications are major drivers of morbidity and mortality, it is unclear to what extent immunological dynamics observed in blood can be translated to the respiratory tract. Indeed, cytokines and antibodies do not seem to correlate between the nasopharynx and peripheral blood during COVID-19 ^12^. Several studies already investigated mucosal immune responses using either bronchoalveolar lavage (BAL) ^13^ or nasopharyngeal and oropharyngeal swabs ^14^ from hospitalized patients, demonstrating increased neutrophil levels and activated alveolar macrophages/monocytes during COVID-19. T cell recruitment to the respiratory tract might be beneficial, as an expansion of CD8^+^ T cell receptor (TCR) clones in BAL has been reported in moderate compared to critical cases ^13^. Another study with severe COVID-19 patients found that increased CD4^+^ T cells in tracheal aspirates associated with survival ^15^. In addition to these findings, several other studies have used nasopharyngeal swabs (NPS) to analyse local responses ^16,17^. However, NPS collect cells only very superficially and mainly provide epithelial and luminal infiltrating cells such as neutrophils and monocytes, while other immune cells, such as T cells are incompletely captured. In contrast, BAL samples and tracheal aspirates provide a clear picture of the lower airways ^18-21^, but are difficult to collect longitudinally, after recovery, or from healthy controls and patients that do not require intubation. Moreover, studies performed on tissue are usually done in severe patients that died from the infection, which creates a bias in the outcomes. As a result, we still have limited understanding of how COVID-19 affects mucosal immunity ^22^. In the current study, we aimed to characterize immune cell dynamics in the upper respiratory tract mucosa during the acute phase and early and later recovery of COVID-19 disease and assess whether alterations persist during convalescence.

To this end, we performed a prospective observational cohort study, for which we recruited patients with PCR-confirmed SARS-CoV-2 infection after hospital admission (Figure 1a). Nasal curettage samples from 20 patients during hospitalization with up to 4 samples per patient were collect and analysed in-depth using mass cytometry. The earliest samples were collected 11 days after symptom onset, which corresponded to 2 days after admission, while the latest sample was collected 82 days after onset (61 days after admission). To address this large temporal range, we stratified hospitalized patients further into those with acute infection (n=9, 2-11 days since admission) or those in early recovery stage (ERS) as defined by having moved from the intensive care unit (ICU) to ward (n=11, 15-61 days since admission, with an ICU stay for a period of 4-55 days). Patient characteristics, co-morbidities, outcome and treatment are shown in Supplementary table 1. Sixteen patients were also sampled 5-6 weeks after hospital discharge (convalescent patients, 42-119 days post symptom onset, with a median of 77 days post onset). In addition, 12 sex and age-matched healthy controls with negative SARS-CoV-2 IgG and no history of symptoms of airway infection were included. In total, 875,564 nasal CD45^+^ immune cells were analysed from a total of 56 samples, comprising of 44 samples collected from 29 COVID-19 patients and 12 samples from healthy donors.

**Figure 1.**
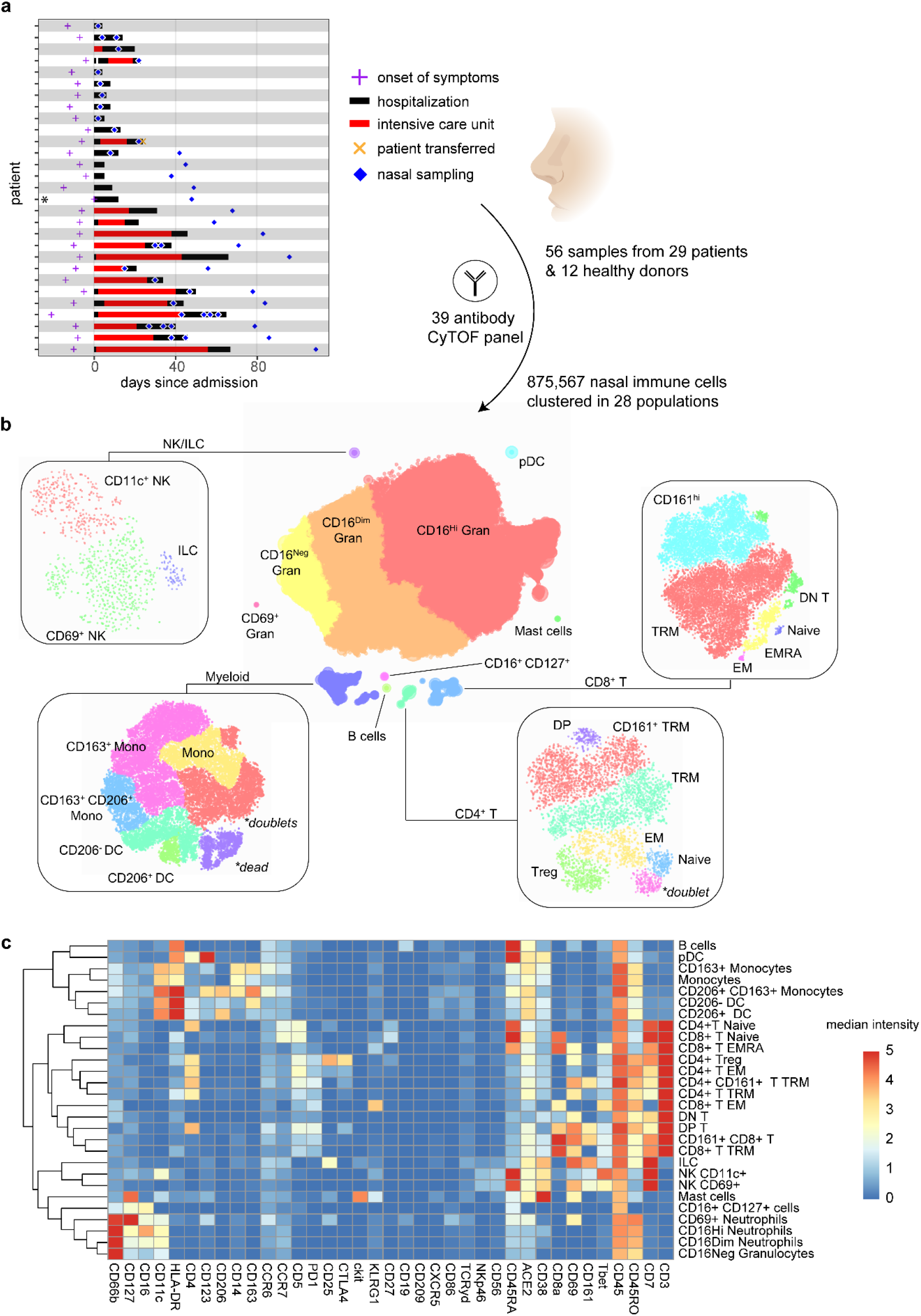
CyTOF analysis of nasal immune cells during and post COVID-19 infection. **a)** Patient timelines. For each included patient, symptom onset (purple cross), hospitalization (black bar), and ICU stay (red bar) are indicated, aligned to the day of admission. The orange x indicates one patient who was included but later transferred to a different hospital. One patient was discharged and then re-admitted a day later. Blue diamonds indicate nasal curettage samples. *one patient was hospitalized for unrelated reasons at time of positive test and symptom onset, hospital admission was set at day of symptom onset. **b)** Hierarchical SNE was used to cluster cellular landmarks on 37 markers, into 12 populations. Some of these populations were then further divided into subpopulations in a second t-SNE embedding at a data level, as indicated in the rectangles. **c)** Heatmap of marker expression per population. Median intensity per population is shown after arcsin transformation. NK = natural killer cells. ILC = innate lymphoid cells, mDC = myeloid dendritic cells, pDC = plasmacytoid dendritic cells, Neutro = neutrophils, Trm = Tissue-resident memory, EM = effector memory, DP = double-positive (CD4+CD8+), DN = double-negative (CD4-CD8-), EMRA = effector memory re-expressing CD45RA. Treg = regulatory T cells.

Immune populations were analysed using a 39-marker mass cytometry panel (Supplementary table 2). Nasal CD45^+^ immune cells were divided into 8 main lineages, which were further sub-clustered into 28 populations (Figure 1b, c). At a lineage level, the nasal immune profile of acute COVID-19 patients was dominated by granulocytes (median 97.2% of all immune cells, IQR: 95.2-97.4%) (Figure 2a). This progressively decreased as granulocyte frequencies were slightly lower in ERS (median 94.8%, IQR: 88.2-97.5%) and even further reduced in convalescent patients (median 78.5%, IQR: 58.8-87.4%), more similar to healthy age-matched donors (median 64%, IQR: 47.3-79.4%). All other lineages, with the exception of monocytes, were decreased during acute infection compared to healthy donors. To understand whether this apparent depletion was related to the increased numbers of granulocytes and monocytes or a true lymphopenia, we performed a normalization step to epithelial cells for each sample. This approach permits an independent assessment of different immune cell populations, while allowing correction for variable sample yield. In healthy donors and recovered patients there was a strong correlation between epithelial and immune cell yields, as expected, while this association was absent in hospitalized patients (Figure 2b). This then suggests that the vastly increased granulocyte numbers within the immune cell analysis are caused by a strong influx of granulocytes into the nasal mucosa of patients. Indeed, when normalizing to epithelial cells, granulocytes and monocytes were highly increased during acute infection and ERS compared to healthy donors (Figure 2c). There was a trend for granulocytes to remain elevated during convalescence compared to the healthy donors (median 0.26 granulocytes per epithelial cell, IQR: 0.09-0.56 in convalescence, versus a median of 0.12, IQR: 0.07-0.20 in healthy donors), although this was not statistically significant (Figure 2c and Supplementary table 3). None of the other main cell lineages: B cells, NK cells, monocytes, pDC, mDC, CD4^+^ T cells and CD8^+^ T cells were statistically different between acute infection and controls. The observation that there is no increase or decrease of total T cell numbers in the nasal mucosa is in contrast with the general T cell depletion that has been observed in peripheral blood ^2,3,6,7,23^.

**Figure 2.**
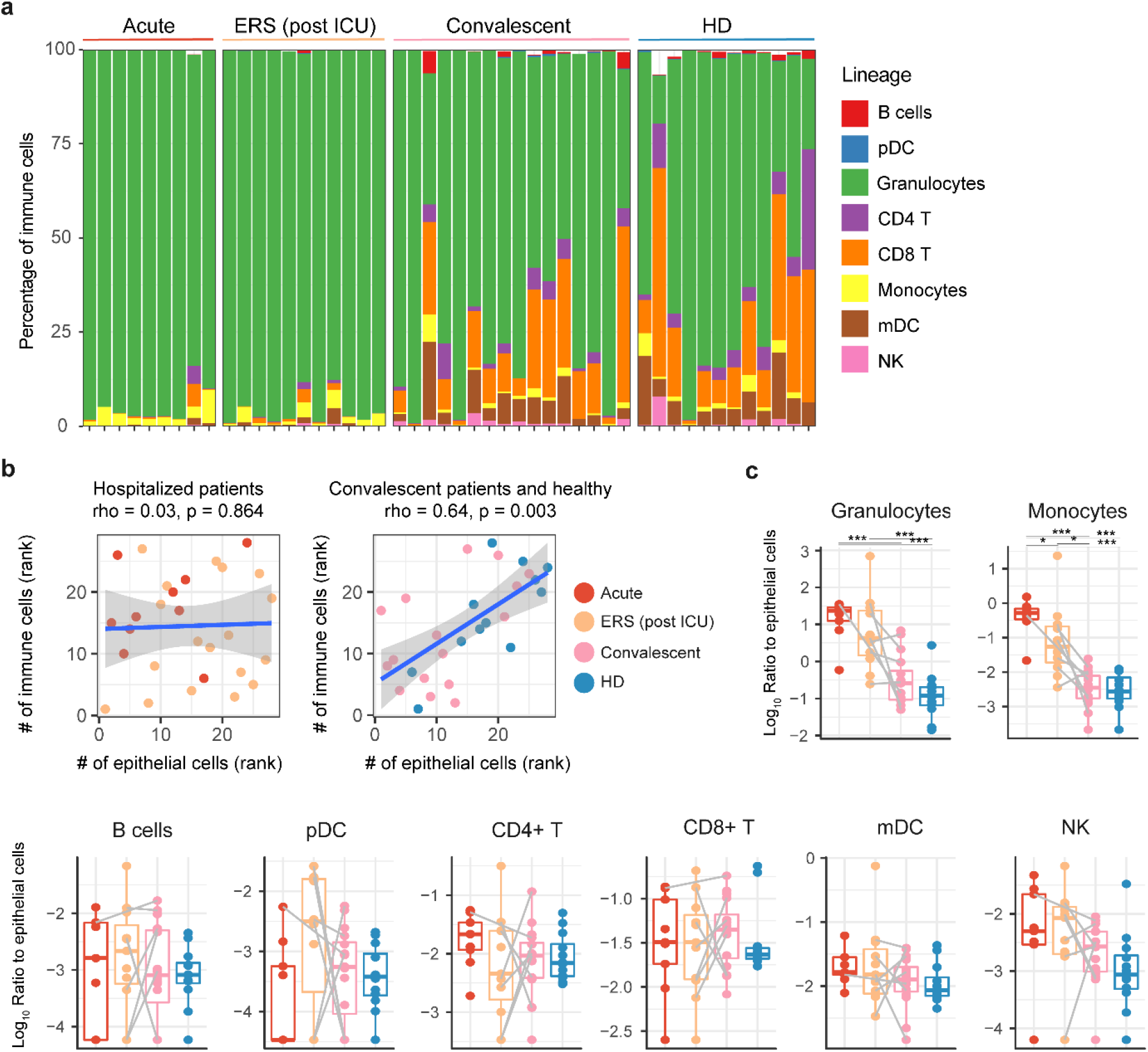
Nasal cell lineage abundance during and post COVID-19. **a)** Stacked bar charts showing the composition of the nasal immune system in acute COVID-19 (red, n=9), during the early recovery phase (post ICU but still in hospital, orange, n = 11), or in COVID-19 convalescence (5-6 weeks post hospital discharge, pink, n=16) or healthy donors (HD, blue, n=12). For patients with multiple samples collected in hospital, only the first sample is shown. **b)** Correlation between nasal immune and epithelial cells for hospitalized patients (left) and convalescent patients and healthy donors (right) are shown. Ranks of individuals are shown with color corresponding to group, as well as regression line and results from spearman correlation analysis. **c)** Ratio of nasal immune cell types normalized to the number of epithelial cells from the same sample. Individuals and boxplots are shown and paired samples between are indicated by grey lines. If a cell type was not detected in at least one sample, half the value of the lowest recorded number was added to each sample, prior to log transformation. pDC = plasmacytoid dendritic cells, mDC = myeloid dendritic cells, NK = natural killer cells. *p<0.05, ***p < 0.001 by linear-mixed model with group as fixed effect and individuals as random effect, with post-hoc testing and Tukey multiple testing correction followed by Benjamini-Hochberg correction for comparing multiple lineages.

On the sub-clustering level (Figure 1b, c), 8 of the 28 defined cell clusters were significantly elevated during various stages of COVID-19, with a clear association with time from hospital admission (Figure 3a,b and Supplementary tables 3 and 4). Three monocyte/macrophage populations were defined, based on expression of CD163 and CD206, and all three were increased during acute infection and ERS compared to convalescent patients and to healthy donors. As expected, subsets of granulocytes were also increased, although with slightly different dynamics during acute infection: CD16^hi^ neutrophils were elevated during acute stage and to a lesser degree in ERS (median of 121.7x and 12.8x increased compared to healthy donors, respectively). CD16^dim^ neutrophils were even more strongly increased during acute infection (median 256.8x) and ERS (median 11.9x). Such CD16^dim^ neutrophils might correspond to activated neutrophils as previously they have been shown to correlate with nasal myeloperoxidase levels and/or they might be related to immature banded neutrophils, recently released from the bone marrow ^24,25^. Furthermore, CD16^neg^ granulocytes, which may in part consist of eosinophils, were increased compared to controls 254.1x and 24.3x during acute stage and ERS, respectively. Although there was no overall change in CD4^+^ T cell numbers, there was an significant increase (median 18.0x) in effector memory (EM, CCR7^-^ CD45RO^+^) CD4^+^ T cells during the acute phase. And although not statistically significant, CD8^+^ T effector memory cells re-expressing CD45RA (EMRA: CCR7^-^ CD45RA^+^) also showed a trend towards increased levels during acute infection compared to healthy donors (median 10.4x increased during acute infection). These findings agree with reports from peripheral blood T cell responses during SARS-CoV-2 infection describing that more specific CD4^+^ T cells are being induced than CD8^+^ T cells, and that the majority of specific CD8^+^ T cells are of a T EMRA phenotype ^26,27^. Levels of these short-lived effector cells returned more or less to numbers observed in healthy donors during ERS and convalescence. Finally, an additional population that was increased in hospitalized patients (both acute and ERS), compared to convalescent stage and healthy donors, were the CD11c^+^ NK cells, which might indicate a NK cell population with increased interferon-producing capacity and effector function ^28^. Thus, dynamic recruitment of various adaptive and innate populations mediating inflammation and antiviral function to the upper respiratory tract was observed during hospitalization with cell type numbers in convalescence that closely resembled levels measured in healthy donors. Of note, we did not observe an increase in nasal B cells in hospitalized patients, which corroborates observations that mucosal antibody levels seem reduced compared to systemic titres in hospitalized patients ^12,29^.

**Figure 3.**
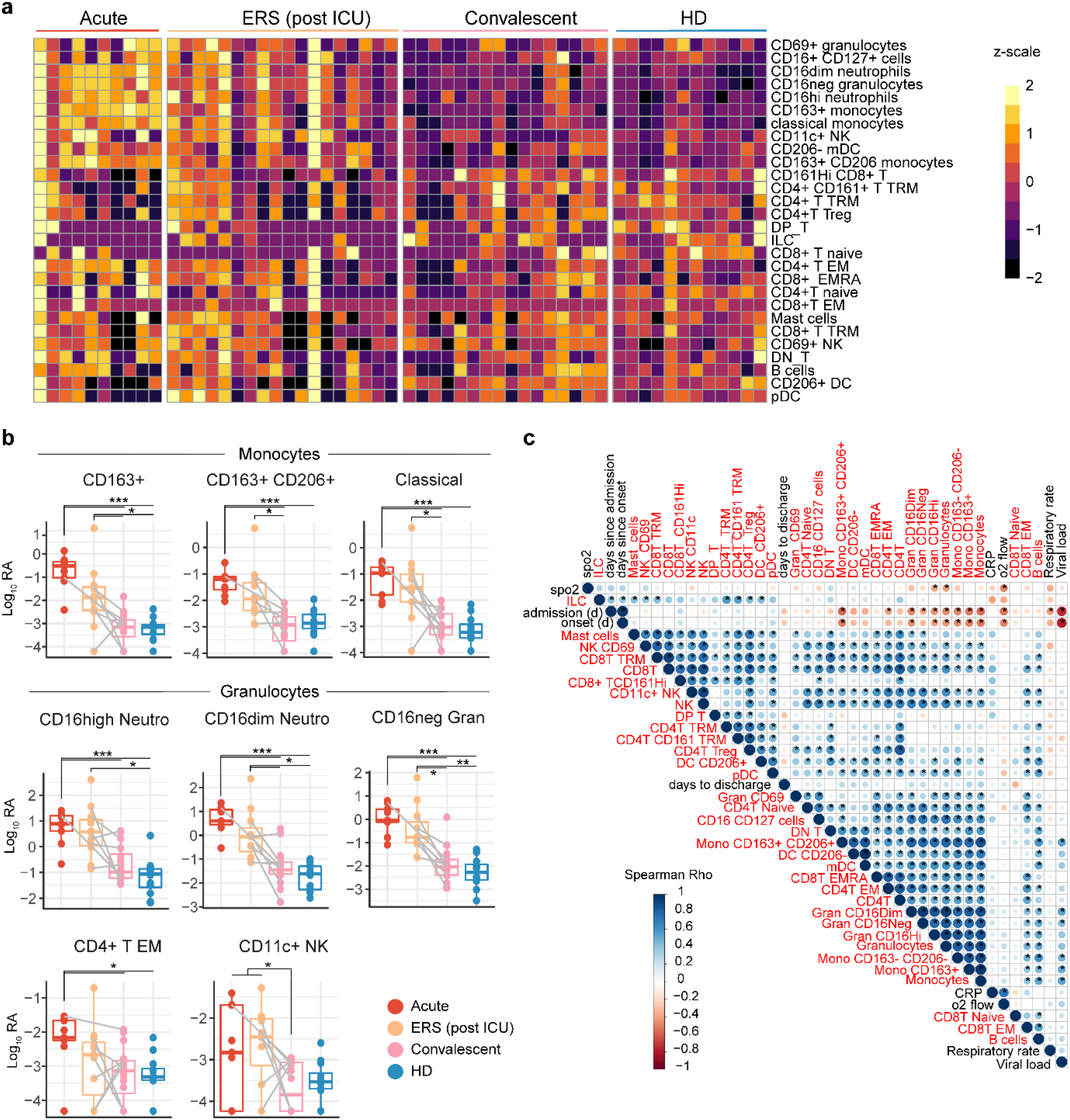
Nasal cell subsets abundance during and post COVID-19. **a**) Heatmap showing log_10_ Relative abundance (RA) of cell clusters. **b**) Boxplots of cell clusters during acute COVID-19 (red, 10 samples from 9 patients), during the early recovery phase (ERS, post ICU but still in hospital, orange, 18 samples from 11 patients n = 11), or in COVID-19 convalescence (5-6 weeks post hospital discharge, pink, n=16) or healthy controls (blue, n=12). Samples are plotted against day of hospital admission, with healthy donors plotted at the right axis separated by a dashed line. *p<0.05, **p<0.01, ***p<0.001, by linear-mixed model with group as fixed effect and individuals as random effect, with post-hoc testing and Tukey multiple testing correction followed by Benjamini-Hochberg correction for comparing multiple subsets. Only first sample per donor in a timepoint (acute or ERS) is shown, but all are included in statistical model. **c)** Correlation plot between nasal immune populations and clinical characteristics. Marginal pairwise correlations were calculated using Spearman’s test between characteristics, using temporally matched data from hospitalized patients. If viral load (VL) data was not available for the time of nasal cell sampling, the closest timepoint with available data was used. VL is calculated as 40-Ct of SARS-CoV-2 PCR, meaning higher values indicate more virus. Rho values are depicted through size and color of symbol. Correlations with a nominal p-value < 0.05 are denoted with an asterisk. Hierarchical clustering based on Euclidian distance of the correlation matrix with complete linkage. Cells are indicated in red, while clinical characteristics are indicated in black.

Next, we analysed whether immune cell types correlated with each other as well as with clinical characteristics during COVID-19 (Figure 3c). Focusing on samples from hospitalized patient, a clear cluster of subsets with positively correlated frequencies was observed, which included monocyte subsets, granulocyte subsets and CD4^+^ T EM and CD8^+^ T EMRA cells. Most of these subsets negatively correlated with days since symptom onset and hospital admission (see also the time-based plots in Supplementary figure 1), indicating that during longer hospitalization the numbers of these cells gradually return to normal levels. Among all cell types, the monocyte subsets, CD16^neg^ granulocytes and CD16^dim^ neutrophils correlated positively with viral load (Figure 3c), suggesting that SARS-CoV-2 virus infection dose-dependently induced nasal recruitment of various immune subsets. We also correlated cell numbers with measurements of disease severity obtained at the same day as nasal sampling, including breathing rate, oxygen saturation and serum CRP concentrations. This revealed that CD16^hi^ neutrophils numbers, as well as total numbers of granulocytes, in nasal mucosa negatively correlated with oxygen saturation levels. As expected, oxygen saturation also negatively associated with applied oxygen flow, but not with viral load. This suggests that fluctuations in viral titers are not related to disease worsening, while the enhanced presence of nasal granulocyte populations is, similar to what has been described for granulocyte numbers in peripheral blood. To understand whether factors like sex, co-morbidities and medication were drivers of nasal immune profiles, we performed multi-dimensional scaling using all cell subsets (Supplementary figure 2). Acute patients clearly clustered separately from healthy donors and convalescent patients, with ERS patients intermediate. Therefore, we visualized covariates separately per group (acute, ERS, convalescent, healthy), showing there was no clear clustering based on any of these covariates, although larger sample sizes might be needed to conclusively exclude any such effects.

Subsequently, we looked more closely at phenotypic expression profiles on the differentially abundant cell clusters and to what extent these profiles normalized after hospital discharge. Although all monocyte subsets significantly increased during acute infection, patients had relatively more CD163^+^ and fewer CD163^+^ CD206^+^ monocytes/macrophages compared to healthy donors, which normalized during recovery (Figure 4a). These CD206^+^ cells are likely fully differentiated tissue-resident macrophages ^30^. CD163^+^ monocytes were also found abundantly in peripheral blood of COVID-19 patients as well as in autopsy lungs and likely represent a recently recruited monocyte population, a hypothesis that was supported by trajectory analysis (Figure 4b) ^15^. Of note, we did not observe non-classical or transitional (CD16^+^) monocytes in the nasal mucosa (Figure 1c). Among all monocyte/macrophage populations, HLA-DR expression was reduced during acute infection and ERS, which normalized in convalescent patients (Figure 4c and Supplementary figure 3). Reduced HLA-DR expression is a characteristic of myeloid-derived suppressor cells, and the expansion of cells in peripheral blood resembling monocyte-derived suppressor cells has been previously reported during severe COVID-19 ^31,32^. Our results suggest that these cells may rapidly seed the upper airway mucosa where they might further differentiate and acquire a macrophage phenotype. CD163^+^ CD206^+^ monocytes/macrophages also expressed elevated levels of the IL-3 receptor CD123 during both acute phase and ERS, while CD163^+^ and CD163^-^ monocyte subsets generally lacked CD123. The expression of CD123 on tissue macrophages has been previously described in patients with histiocytic necrotizing lymphadenitis^33^. The functional consequences of this upregulation of the IL3R on nasal macrophages during COVID-19 remain to be elucidated, but this finding further supports the importance of the IL-3/IL3-R axis in COVID-19 ^34^.

**Figure 4.**
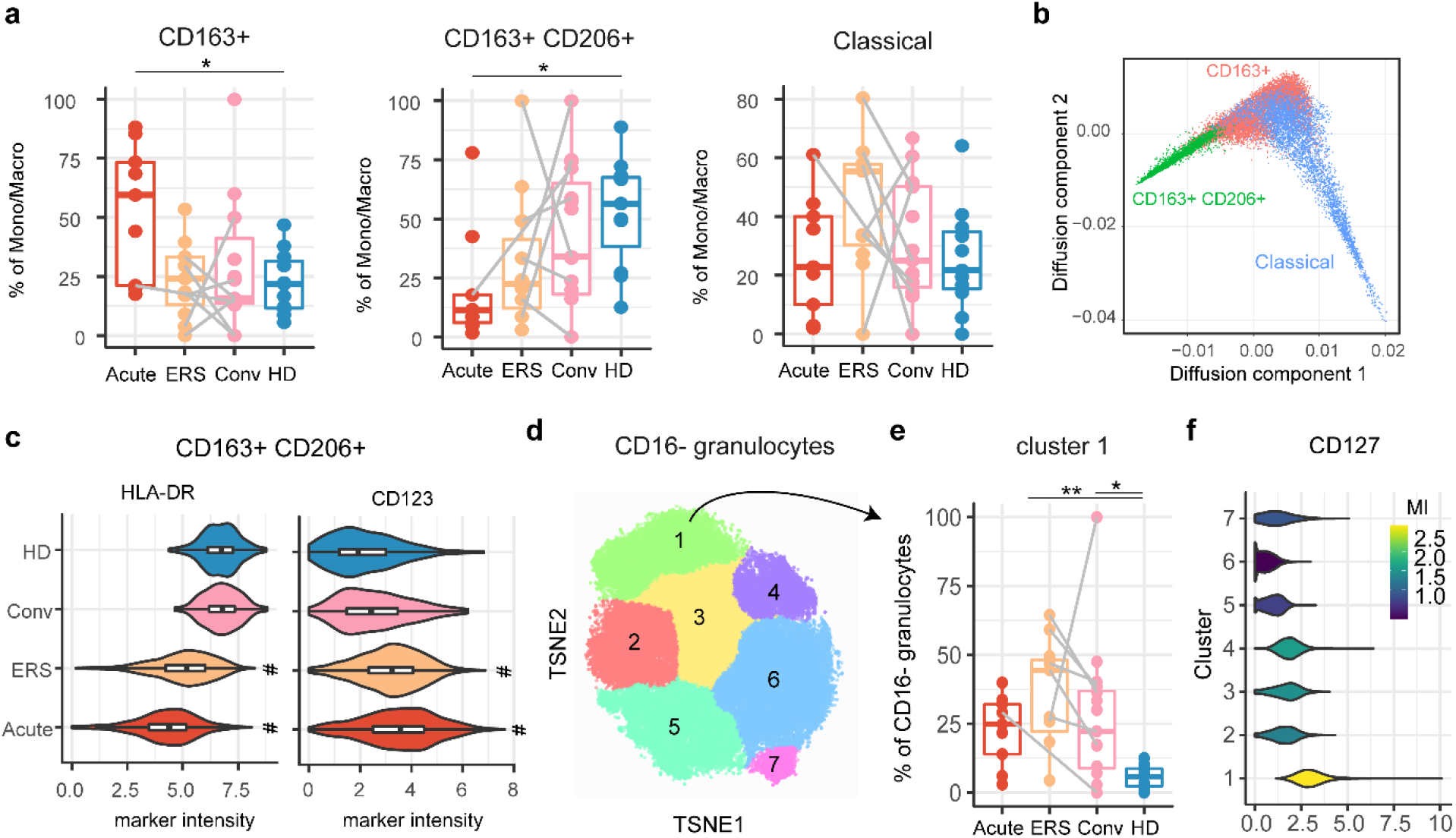
Phenotypic changes of innate and adaptive cell subsets during and post COVID-19. **a)** Percentages of monocytes expressing CD163, CD163 and CD206 or neither (Classical). Subsets frequencies are shown for during acute COVID-19 (red, n=9 samples), during the early recovery phase (post ICU but still in hospital, orange, n = 11), or in COVID-19 convalescence (5-6 weeks post hospital discharge, pink, n=16) or healthy donors (blue, n=12). Boxplots and individual datapoints are depicted with paired patient samples connected by a grey line. Only the first sample per patient for a given timepoint/group is shown, although all are used for statistical modeling. * p < 0.05, p-values by linear-mixed model with post-hoc testing and Tukey multiple testing correction. HD = healthy donor, Conv = convalescent. **b)** Diffusion map showing trajectory analysis of monocyte/macrophage subsets. All cells are colored according to their clustered phenotype. **c)** Violin plots showing expression of HLA-DR and CD123 on CD163+ CD206+ monocytes/macrophages. ^#^p<0.001 compared to healthy donors and convalescent patients, using pairwise Wilcoxon test and Bonferroni correction for multiple testing. **d)** Clustering of CD16-granulocytes using tSNE and Gaussian mean shift. **e)** Proportion of cluster 1 within CD16-granulocytes. * p < 0.05, ** p < 0.01, p-values by linear-mixed model with post-hoc testing and Tukey multiple testing correction. **f)** Violin plots showing expression of CD127 per cluster.

We then investigated more closely CD16^-^ granulocytes, by further grouping them into 7 sub-clusters (Figure 4d). Sub-cluster 1, characterized by increased CD127 expression, was significantly increased in patients during hospitalization (median 28.3%) but also during convalescence (median 22.2%) compared to healthy donors (median 5.8%, Figure 4e,f). It has been shown that engagement of IL-7 with its receptor CD127 on eosinophils leads to increased survival and activation of eosinophils; and in airway allergen challenge in allergic asthmatics, IL-7 levels in BAL strongly correlated with eosinophils ^35^. Thus while total numbers of granulocytes returned to similar levels as observed in healthy donors, alterations in their phenotype, and possibly their functionality, remained visible during convalescence. The half-life of granulocytes is in the region of hours to days, suggesting either an ongoing recruitment of altered cells or the continued local perturbation after or during entering the respiratory mucosa ^35^.

The subset of CD4^+^ EM T cells was also increased during acute infection. To investigate their phenotype more closely, we analysed the expression of markers of activation (CD38, HLA-DR), exhaustion (PD1) or inhibition (CTLA-4) on these cells (Figure 5a). CD38 and CTLA4 were increased during the acute phase, but normalized at later timepoints, while HLA-DR and PD1 remained expressed at a higher level even during convalescence as compared to healthy donors. This induction of regulatory and inhibitory markers mirrors what has been described in blood and likely reflects attempts of the immune system to restrain excessive activation. Finally, we aimed to assess whether long-term protective CD8^+^ T cell immunity develops in nasal mucosa to serve as gatekeepers against re-infections. Mouse models have shown that nasal CD8^+^ tissue-resident memory (TRM) T cells specific for influenza persisted in nasal mucosa following infection and efficiently controlled secondary infections ^36^. The majority of nasal CD8^+^ T cells in our samples highly expressed CD69, which is used to define resident-memory T cells ^37^, while very few naïve CD8^+^ T cells were present. Although it is possible that some activated CD8^+^ T cells upregulate CD69 without being true TRM, the cluster of CD8^+^ TRM expressed very little KLRG1, which is congruent with a TRM phenotype (Figure 1c) ^38^. Sub-clustering of CD8^+^ TRM showed variable expression of activation markers CD38, HLA-DR and Tbet (Figure 5b). Within this embedding, CD8^+^ TRM from acute phase, ERS, convalescent patients and controls clustered differentially, indicative of altered phenotypes during and following COVID-19 (Figure 5c). Indeed, sub-cluster frequencies significantly differed between the groups (Figure 5d, e). Sub-cluster 5, marked by expression of HLA-DR, Tbet and CD38, was increased in hospitalized patients, while sub-cluster 3, expressing only HLA-DR and CD38, was higher in convalescent patients compared to healthy donors, as were all CD38^+^ TRMs. Thus CD8^+^ TRM had an increased activation profile, which persisted at least 5-6 weeks after hospital discharge. To demonstrate antigen specificity, we then attempted to compare the T cell receptor (TCR) repertoire in nasal samples from convalescent patients with SARS-CoV-2 reactive CD8^+^ and CD4^+^ T cells isolated from paired peripheral blood (Supplementary figure 4). For one severe, convalescent patient we obtained >10 unique TCRs from both nasal cells and sorted SARS-CoV-2 specific peripheral blood cells (Supplementary figure 5). The TCR repertoire in the nasal mucosa of this patient was broad (218 unique TCRs), including one dominant TCR accounting for 12.2% of all TCR reads. This TCR clone was also present in paired peripheral blood SARS-CoV-2 specific CD8^+^ T cells (4% of reads), but not in sorted CD4^+^ T cells (Figure 5f). As this sample was collected 2 months after viral clearance and 80.4% of the nasal CD8^+^ T cells for this patient were of a TRM phenotype (Figure 5g), this indicates that antigen-specific tissue-resident memory was induced. Of note, the number of unique SARS-CoV-2 specific TCRs detected and their overlap between nose and peripheral blood might be underestimated as blood CD8^+^ T cells were isolated based on reactivity towards structural proteins, while CD8^+^ T cell reactivity is also directed against non-structural proteins in severe COVID-19 patients ^39-41^. Taken together, we demonstrated SARS-CoV-2 specific CD8^+^ T cells can persist in the nasal mucosa months after viral clearance, based on their activated phenotype and overlapping TCR clonotype with SARS-CoV-2 reactive CD8^+^ T cells in blood. This is suggestive of the establishment of local protective immune memory responses that could rapidly control and attenuate re-infections by SARS-CoV-2.

**Figure 5.**
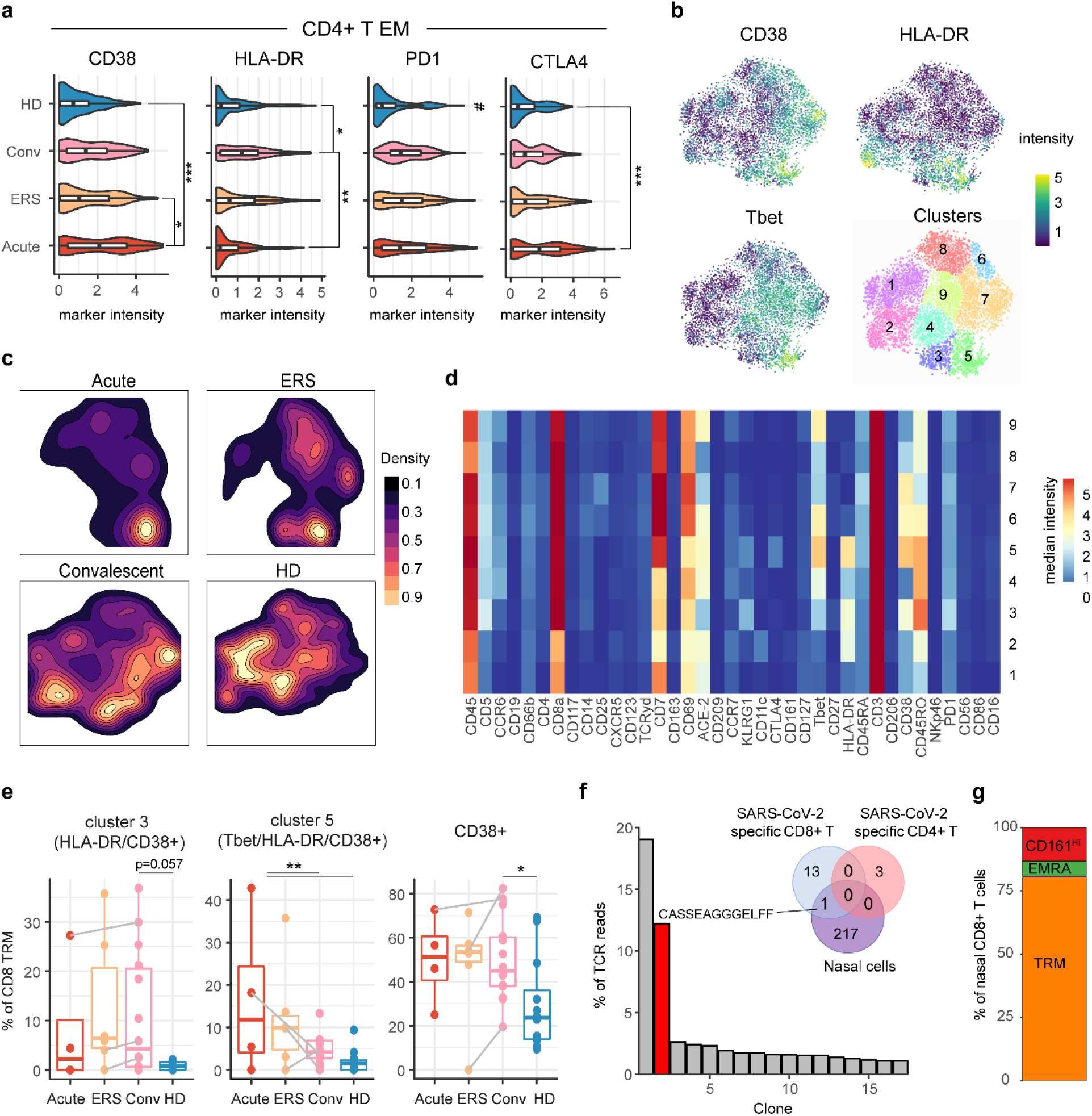
Activated SARS-CoV-2 specific T cells are present in the nasal mucosa after recovery. **a)** Phenotype of CD4+ T effector memory (EM) cells. Intensity for four activation/exhaustion/regulatory markers are shown as violin plots, with box plots inlayed. **p<0.01, ***p<0.001, ^#^p<0.001 compared to all other groups, using pairwise Wilcoxon test and Bonferroni correction for multiple testing. **b**) tSNE analysis of CD8+ TRM cells. Expression of CD38, HLA-DR or Tbet overlaid onto tSNE embedding. Bottom-right, clustering of tSNE using Gaussian mean shift, with clusters overlaid onto the embedding. Clusters numbers are indicated. **c)** Two-dimensional kernel density estimation from tSNE embedding of cells belonging to the 4 groups group. **d)** Heatmap showing median expression for all markers per CD8^+^ T cell cluster. **e)** Percentage of CD8+ TRM belonging to either cluster 3 or cluster 5, or all clusters expressing CD38. * p < 0.05, ** p < 0.01, p-values by linear-mixed model with post-hoc testing and Tukey multiple testing correction. **f)** Bar plots showing the frequency of TCR clonotypes in nasal samples collected from one severe donor, only clones with frequency >1% are shown. Red colored bar indicates the clone that was also found in sorted SARS-CoV-2 specific CD8^+^ T cells. Venn diagram shows the overlap between TCR clones from nasal sample and paired SARS-CoV-2 specific CD4^+^ T cells or CD8^+^ T cells in PBMC. **g**) Bar plot showing nasal CD8^+^ T cell composition for patient with dominant SARS-CoV-2 specific clone. Naive and EM cells were too infrequent to be visible and are not labeled for this sample.

In conclusion, we provide the first comprehensive overview to date of how COVID-19 affects the nasal mucosal immune system during acute infection and during early and later recovery stages. Acute COVID-19 led to transient increases in granulocytes and monocyte subsets, a NK cell subset and CD4^+^ T EM. Elements of this strong pro-inflammatory response positively correlated with viral load and nasal neutrophil numbers in addition correlated negatively with oxygen saturation in blood. We show several similarities with reports from blood, including a strong CD4^+^ T EM cell response marked by markers of activation, but also exhaustion and inhibition, as well as the presence of HLA-DR^low^ monocyte subsets in the nasal mucosa. A striking difference from studies characterizing blood was the absence of a general lymphopenia in the nasal mucosa. We also provided further characterization of cellular subsets infiltrating the mucosa during disease, such as CD11c^+^ NK cells, CD123-expressing differentiated macrophages and CD127 expression on CD16^-^ granulocytes. During early and later recovery stages of disease most of the cell numbers progressively returned to levels comparable to age-matched healthy donors. However, several phenotypic changes in nasal immune populations persisted even during recovery. For example CD127-expressing granulocytes remained elevated during convalescence, while also residual increased activation of CD4^+^ and CD8^+^ T cells after hospital discharge was observed. Moreover, by comparing nasal TCR clonotypes with antigen-specific cells from blood, we were able to demonstrate that SARS-CoV-2 specific CD8^+^ T cells can seed the nasal tissue and these tissue-resident memory cells can persist at least up to 2 months post viral clearance. Future studies should focus on mucosal immune responses in SARS-CoV-2 infected people with only mild symptoms to provide insights into which mucosal immune populations associate with control of infection. Moreover, the collection of samples very early in infection before hospital admission would allow to stratify and predict disease course. Likewise, this study highlights the need for studying patients post infection to understand persistent immunological perturbations following SARS-CoV-2 infection. Altogether, this study provides unique insights into the mucosal immune cell dynamics both during acute and recovery of COVID-19.

## Data Availability

Raw data is available upon reasonable request from corresponding author under compliance with ethical and regulatory approvals.

## Acknowledgments

We thank all patients and healthy volunteers for taking part in this study. This work was supported by a MKMD-COVID-19 grant (no. 114025007) from ZonMW and Proefdiervrij. This work was also supported by Wake Up To Corona crowdfunding by Leids Universitair Fonds. SPJ is supported by a LUMC Gisela Thier Fellowship. Figure 1 was partly made with BioRender (https://biorender.com/). The authors gratefully acknowledge the Flow cytometry Core Facility (FCF) at LUMC, Leiden, Netherlands (https://www.lumc.nl/research/facilities/fcf), coordinated by dr. K. Schepers and M. Hameetman, run by the FCF Operators E.F.E de Haas, J.P. Jansen, D.M. Lowie, S. van de Pas, and G.IJ. Reyneveld (Directors: Prof. F.J.T. Staal and Prof. J.J.M. van Dongen) for technical support in the mass cytometry studies.

The authors declare no competing interests.

## Methods

### Study design and ethics

In this prospective observational cohort study, adult patients with PCR-confirmed COVID-19 who were admitted to our academic hospital were recruited. All hospitalized patients had hypoxia. The study was performed at the Leiden University Medical Center, Leiden, the Netherlands, from patients included from April 2020 until December 2020. After informed consent was obtained, longitudinal sampling was performed for the duration of the hospital admission, and one convalescent sample was obtained at the outpatient follow-up appointment, which was scheduled six weeks after hospital discharge. Ethical approval was obtained from the Medical Ethical Committee Leiden-Den Haag-Delft (NL73740.058.20). The trial was registered in the Dutch Trial Registry (NL8589). As ICU patients had substantial breathing support, we were unable to collect nasal mucosal cells from patients on the ICU. Twelve healthy donors were included in the study. These were all sixty years or older, and with a male:female ratio of 2:1, in order to match the patient population. The healthy donors had no recent history of symptoms of airway infection (fever, cough, hypoxia, rhinorrhea, myalgia, anosmia and or ageusia or fatigue), and were included after confirmed negative SARS-CoV-2 IgG.

### Nasal cell collection and storage

Nasal cells were collected by gently scraping the nasal inferior turbinate using curettes (Rhino-Pro®, Arlington Scientific), as described previously^42^, and placing them in a tube containing pre-cooled 8mL sterile PBS containing 5mM EDTA (Life Technologies). Such samples provide immune cells from the mucosa that are not found in the lumen (including lymphoid subsets) ^42^, and have been used previously to study nasal immune responses during controlled viral and bacterial infections ^43,44^. Per patient and timepoint, two curettes from one nostril were collected. Cells were dislodged by pipetting liquid up and down the tip of curette and spun down at 300xg for 10’ at 4°C. Supernatant was completely removed and cells were resuspended in 500µL of PBS. For fixation, an equal amount of freshly prepared 8% formaldehyde (Fisher Scientific) was then added, followed by 30 minutes incubation at room temperature. Cells were then spun down at 800xg for 10’. The supernatant was completely removed and the pellet resuspended in 1mL heat-inactivated fetal bovine serum containing 10% DMSO and moved to a cryovial. Cryovials were frozen in a Mr. Frosty Frosty™ freezing container (ThermoFisher Scientific) at -80°C and moved to liquid nitrogen within three days.

### CyTOF staining

Nasal samples were barcoded and measured in batches. In every batch, one aliquot of PBMCs from a reference sample was included to be able to normalize staining between batches. First, cells were thawed in 2mL RPMI + 50% FBS and spun down for 10’ at 1600rpm at room temperature. Supernatant was discarded by pipetting. Reference PBMC were washed with 2mL of PBS and then fixed with 4% formaldehyde for 15’ at room temperature. Reference PBMC were washed 2x with 2mL BD Perm/Wash (BD). Nasal cells were washed 1x with 1mL BD Perm/Wash, and if clumps were visible, cells were filtered over a 100µm filter (ThermoFisher Scientific). Nasal cells and reference PBMCs were resuspended in 50µL Perm/Wash and then 50µL barcode mix targeting β2 microglobulin (B2M) was added to each individual sample in a 6-choose-3 scheme using Cadmiums 106, 110, 111, 112, 114 and 116 ^45,46^. Samples were incubated for 30’ at room temperature and then washed with 4mL Cell Staining Buffer (Fluidigm). Cells were spun 5’ at 800xg, supernatant removed and resuspended and combined into 3mL of Perm/Wash. Cells were spun again 5’ at 800xg and were resuspended in 45µL Perm/Wash. FcR block (Biolegend, 5µL) and heparin (0.5µL, 100U/mL) were added to prevent aspecific binding of antibodies and cells were incubated for 20’ at room temperature^47^. Then 50µL of antibody cocktail (Table S1) was added, followed by a 45’ incubation at room temperature. Cells were then washed twice with 2mL Cell Staining Buffer and spun down for 5’ at 800xg. DNA was then stained overnight at 4°C using 1mL Fix and perm buffer (Fluidigm) containing 1000x diluted Intercalator-Ir (Fluidigm). Cells were then washed with Cell Staining Buffer, counted and divided into tubes of 1×10^6^ cells and pelleted down. Tubes were then washed and resuspended in cell acquisition solution (CAS, Fluidigm) with EQ Four Element Calibration Beads (Fluidigm) and acquired on a Helios mass cytometer (Fluidigm) at the Flow cytometry Core Facility (FCF) of Leiden University Medical Center (LUMC) in Leiden, Netherlands (https://www.lumc.nl/research/facilities/fcf).

### Data preprocessing and clustering

An outline of data pre-processing steps is shown in Supplementary figure 6. Debris and normalization beads were filtered from .FCS files using the ‘CyTOFclean’ package (v1.0.1) ^48^. Single cells were then manually gated based on DNA stain and the ‘CATALYST’ package (v1.12.2) and single-stain controls were used to compensate data using the non-negative linear least squares method ^49^. One by one plots were used to confirm correct compensation of data. Then epithelial and immune cells were manually gated based on CD45 and EpCAM expression, with exclusion of cPARP positive apoptotic cells, as well as immune doublets (CD14+CD3+, CD66b+CD3+, CD14+CD66b+). Subsequently, ‘CATALYST’ package (v1.12.2) was used to debarcode immune and epithelial cells individually per batch. FCS files were then normalized using the reference PBMCs and the CyTOFBatchAdjust package with 99 percentile scaling for each marker individually ^50^. The marker CD69 was not present in the reference PBMC at sufficient levels to scale and was thus not normalized. Signal intensity and clustering of reference samples before and after normalization was used to verify appropriate normalization. Clustering of cells into populations was done using hierarchical stochastic neighbor embedding (hSNE) or tSNE with Cytosplore software (v2.3.0, https://www.cytosplore.org/), using all markers minus EpCAM and cPARP ^51^. All t-SNE analyses were performed with complexity = 30. Diffusion map of monocytes was created using ‘destiny’ package (v.3.2.0) with k = 1000, using the markers: HLA-DR, CD11c, CD163, ACE-2, CD45RO, CD14, CD38, CD127, CD206, CD86, CD4, CD123 and CD45RA.

### SARS-CoV-2 reactive T-cell isolation

PBMCs from convalescent COVID-19 patients were isolated from fresh whole blood using Ficoll-Isopaque and cryopreserved until further use. PBMCs were thawed and immediately used for overnight stimulation assay. For the stimulation assay, 1×10 PBMCs were seeded in 100µL IMDM (Lonza) + 10% FCS (Sigma) + 1,4% L-glutamine (Lonza) + 1% Pen/Strep (Lonza) and in the presence of 1µg/mL SARS-CoV-2 peptide pool or 1% DMSO (negative control). The SARS-CoV-2 peptide pool consisted of 15-mer peptides derived from nucleocapsid (N) (Miltenyi, cat#130-126-699), membrane (M) (Miltenyi, cat#130-126-703) and most immunogenic sequences from the spike protein (S) (Miltenyi, cat#130-126-701). Peptides were dissolved and used according to manufacturer’s protocol. After 16 hours the PBMCs were washed and stained for CD4-FITC (BD, cat#555346), CD8-PeCy7 (BD, cat#557746), CD154-Pacific Blue (Biolegend, cat#310820) and CD137-APC (BD, cat#550890) for 30 minutes at 4°C. PBMCs were washed, stained and sorted in phenol red free DMEM (Gibco) + 2% FCS + 1% Pen/Strep.

### TCR identification

TCRβ sequences were identified as previously described with minor modifications ^52^. In brief, RNA was isolated from 1×10^2^ to 1×10^6^ cells using the ReliaPrep RNA cell Miniprep system (Promega). cDNA was generated using an Oligo dT-I.S ^53^, SMARTScribe Reverse Transcriptase (Takara, Clontech), and a SA.rt template switching oligo forward primer. If needed the complete cDNA was pre-amplified for 18 cycles with I.S. primers that anneal both to the SA.rt and oligo dT IS region. Barcoded TCR PCR product was generated in two rounds of PCR. In the first PCR, TCRβ product was generated, in a second PCR the first PCR product was used to include a 2-sided barcode sequence that allows discrimination between TCRs of different T cell populations. PCR products of different T cell populations were pooled after which TCR sequences were identified by NovaSeq (GenomeScan). NovaSeq data were analysed using MiXCR software to determine the Vβ family and CDR3 regions. CDR3 regions were analysed in Rstudio and CDR3 sequences with ≤50 reads, that were non-functional or occurred on all samples were excluded from the analysis.

## Statistical analysis

Statistical differences in cellular abundance between groups were compared with a linear mixed model, in which individual patients were included as random effect and groups (acute, ERS, convalescent and healthy donors) as fixed effect, using the ‘lme4’ (v.1.1-23) and ‘lmerTest’ (v3.1-2) packages. Post-hoc comparison of the groups included in the model was conducted with the ‘emmeans’ package (v1.4.8), using Tukey correction. For the comparison of multiple subsets or lineages, Benjamini-Hochberg multiple-testing correction was used. Correlation values were calculated with Spearman non-parametric tests. Marker expression between groups was compared with Wilcoxon test, with Bonferroni correction for multiple markers and groups as in Seurat ^54^. Statistical tests were performed two-tailed. All analyses were performed with R version 4.0.1, except for the ‘cytofclean’ package done in R3.6.3, using Rstudio (v1.2.5033)

