## Supplementary tables and figures for "Prolonged activation of nasal immune cell populations and development of tissue-resident SARS-CoV-2 specific CD8 T cell responses following COVID-19"

***Supplementary Table 1.*** *Patient and control characteristics. ND = not determined. NA = not applicable.*

| | **Patients (n=29)** | **Healthy donors (n=12)** |
|---|---|---|
| Female sex (%) | 9 (31.0) | 4 (33.3) |
| Median age (range) | 62 (19-78) | 64 (60-72) |
| Median BMI (min-max) | 28.1 (17.6-42.3) | ND |
| Disease course | | NA |
| Median days of hospital stay (range) | 21 (1-67) | |
| ICU stay | 16 | |
| Live discharge | 28 | |
| Transfer to other hospital | 1 | |
| Comorbidities | | ND |
| Chronic cardiac disease | 6 | |
| Hypertension | 6 | |
| Chronic obstructive pulmonary disease | 2 | |
| Asthma | 4 | |
| Chronic kidney disease | 1 | |
| Chronic liver disease | 2 | |
| Chronic neurological disorder | 2 | |
| Diabetes | 10 | |
| Malignant neoplasm | 6 | |
| Thromboembolic event | 6 | |
| Coagulopathy | 1 | |
| Medication | | ND |
| Azathioprine | 1 | |
| Beclomethasone | 2 | |
| Chloroquine | 1 | |
| Dexamethasone | 7 | |
| Fluticasone | 3 | |
| Hydrochlorothiazide | 1 | |
| Hydrocortisone | 2 | |
| Hydroxychloroquine | 3 | |
| Metamizole | 2 | |
| Prednisolone | 3 | |

***Supplementary Table 2.*** *List of antibodies used for mass cytometry*

| Label | Specificity | Clone | Antibody vendor | Catalogue number | Dilution |
|---|---|---|---|---|---|
| $^{89}$Y | CD45 | HI30 | Fluidigm | 3089003B | 1/200 |
| $^{115}$In | CD5 | UCHT2 | Biolegend | 300602 | 1/100 |
| $^{141}$Pr | CD196 (CCR6) | G034E3 | Fluidigm | 3141003A | 1/200 |
| $^{142}$Nd | CD19 | HIB19 | Fluidigm | 3142001B | 1/200 |
| $^{143}$Nd | cPARP | F21-852 | Fluidigm | 3143011A | 1/100 |
| $^{144}$Nd | CD66b | REA306 | Miltenyi Biotech | 130-108-019 | 1/50 |
| $^{145}$Nd | CD4 | RPA-T4 | Biolegend | 300541 | 1/100 |
| $^{146}$Nd | CD8a | RPA-T8 | Biolegend | 301053 | 1/200 |
| $^{147}$Sm | CD117 (C-kit) | 104D2 | BioLegend | 313202 | 1/100 |
| $^{148}$Nd | CD14 | M5E2 | Biolegend | 301843 | 1/100 |
| $^{149}$Sm | CD25 (IL-2Ra) | 2A3 | Fluidigm | 3149010B | 1/100 |
| $^{150}$Nd | CD185 (CXCR5) | J252D4 | BioLegend | 356902 | 1/100 |
| $^{151}$Eu | CD123 | 6H6 | Fluidigm | 3151001B | 1/100 |
| $^{152}$Sm | TCRγδ | 11F2 | Fluidigm | 3152008B | 1/50 |
| $^{153}$Eu | CD7 | CD7-6B7 | Biolegend | 343111 | 1/100 |
| $^{154}$Sm | CD163 | GHI/61 | Biolegend | 333602 | 1/100 |
| $^{155}$Gd | CD69 | FN50 | BioLegend | 310939 | 1/200 |
| $^{157}$Gd | ACE-2 | AC18F | Novus Biologicals | NBP2-80035-100UG | 1/100 |
| $^{158}$Gd | CD209 | 9E9A8 | Biolegend | 330102 | 1/100 |
| $^{159}$Tb | CD197 (CCR7) | G043H7 | Biolegend | 353237 | 1/200 |
| $^{160}$Gd | Epcam | AUA1 | Invitrogen | MA5-13917 | 1/50 |
| $^{161}$Dy | KLRG1 (MAFA) | REA261 | Miltenyi Biotech | Special order | 1/100 |
| $^{162}$Dy | CD11c | Bu15 | Biolegend | 337221 | 1/200 |
| $^{163}$Dy | CD152 (CTLA-4) | BNI3 | BioLegend | 369602 | 1/100 |
| $^{164}$Dy | CD161 | HP-3G10 | Biolegend | 339919 | 1/100 |
| $^{165}$Ho | CD127 (IL-7Ra) | AO19D5 | Fluidigm | 3165008B | 1/200 |
| $^{166}$Er | Tbet | 4B10 | BioLegend | 644825 | 1/100 |
| $^{167}$Er | CD27 | O323 | Biolegend | 302839 | 1/200 |
| $^{168}$Er | HLA-DR | L243 | Biolegend | 307651 | 1/200 |
| $^{169}$Tm | CD45RA | HI100 | Fluidigm | 3169008B | 1/100 |
| $^{170}$Er | CD3 | UCHT1 | Biolegend | 300443 | 1/100 |
| $^{171}$Yb | CD206 | 15-2 | Biolegend | 321127 | 1/200 |
| $^{172}$Yb | CD38 | HIT2 | Biolegend | 303535 | 1/200 |
| $^{173}$Yb | CD45RO | UCHL1 | Biolegend | 304239 | 1/100 |
| $^{174}$Yb | CD335 (NKp46) | 92E | Biolegend | 331902 | 1/100 |
| $^{175}$Yb | PD-1 | EH12.2H7 | Biolegend | 329941 | 1/100 |
| $^{176}$Yb | CD56 | B159/NCAM16.2 | BD Biosciences | 555514 | 1/100 |
| $^{198}$Pt | CD86 | IT2.2 | BioLegend | 305435 | 1/200 |
| $^{209}$BI | CD16 | 3G8 | Fluidigm | 3209002B | 1/200 |
| $^{106}$Cd | B2M | 2M2 | BioLegend | 316302 | 1/50 |
| $^{110}$Cd | B2M | 2M2 | BioLegend | 316302 | 1/50 |
| $^{111}$Cd | B2M | 2M2 | BioLegend | 316302 | 1/50 |
| $^{112}$Cd | B2M | 2M2 | BioLegend | 316302 | 1/50 |
| $^{114}$Cd | B2M | 2M2 | BioLegend | 316302 | 1/50 |
| $^{116}$Cd | B2M | 2M2 | BioLegend | 316302 | 1/50 |

***Supplementary table 3. See supplementary Excel file****. Results from statistical model comparing 3 groups: all hospitalized patients, convalescence stage 5-6 weeks post discharge, and healthy donors. Results are from linear mixed model, using log-transformed cell numbers divided by epithelial cells, using groups as fixed effects and donors as random effects to account for repeated sampling. Healthy donors were used as base group. Estimates and standard errors are from log10-transformed values. P-values are calculated using post-hoc testing with 'emmeans' package in R, followed by Tukey multiple testing correction. Both uncorrected and Benjamini-Hochberg corrected p-values for comparing 8 lineages or 26 subsets are given. Populations that are lineages are indicated in yellow. Group estimates above or below 0.5 log difference from healthy donors are highlighted, as well as p-values below 0.05.*

***Supplementary table 4. See supplementary Excel file****. Results from statistical model comparing the 4 groups (acute hospitalization, post ICU/early recovery stage, convalescence stage 5-6 weeks post discharge, and healthy donors. Results are from linear mixed model, using log-transformed cell numbers divided by epithelial cells, using groups as fixed effects and donors as random effects to account for repeated sampling. Healthy donors were used as base group. Estimates and standard errors are from log10-transformed values. P-values are calculated using post-hoc testing with 'emmeans' package in R, followed by Tukey multiple testing correction. Both uncorrected and Benjamini-Hochberg corrected p-values for comparing 8 lineages or 26 subsets are given. Populations that are lineages are indicated in yellow. Group estimates above or below 0.5 log difference from healthy donors are highlighted, as well as p-values below 0.05.*

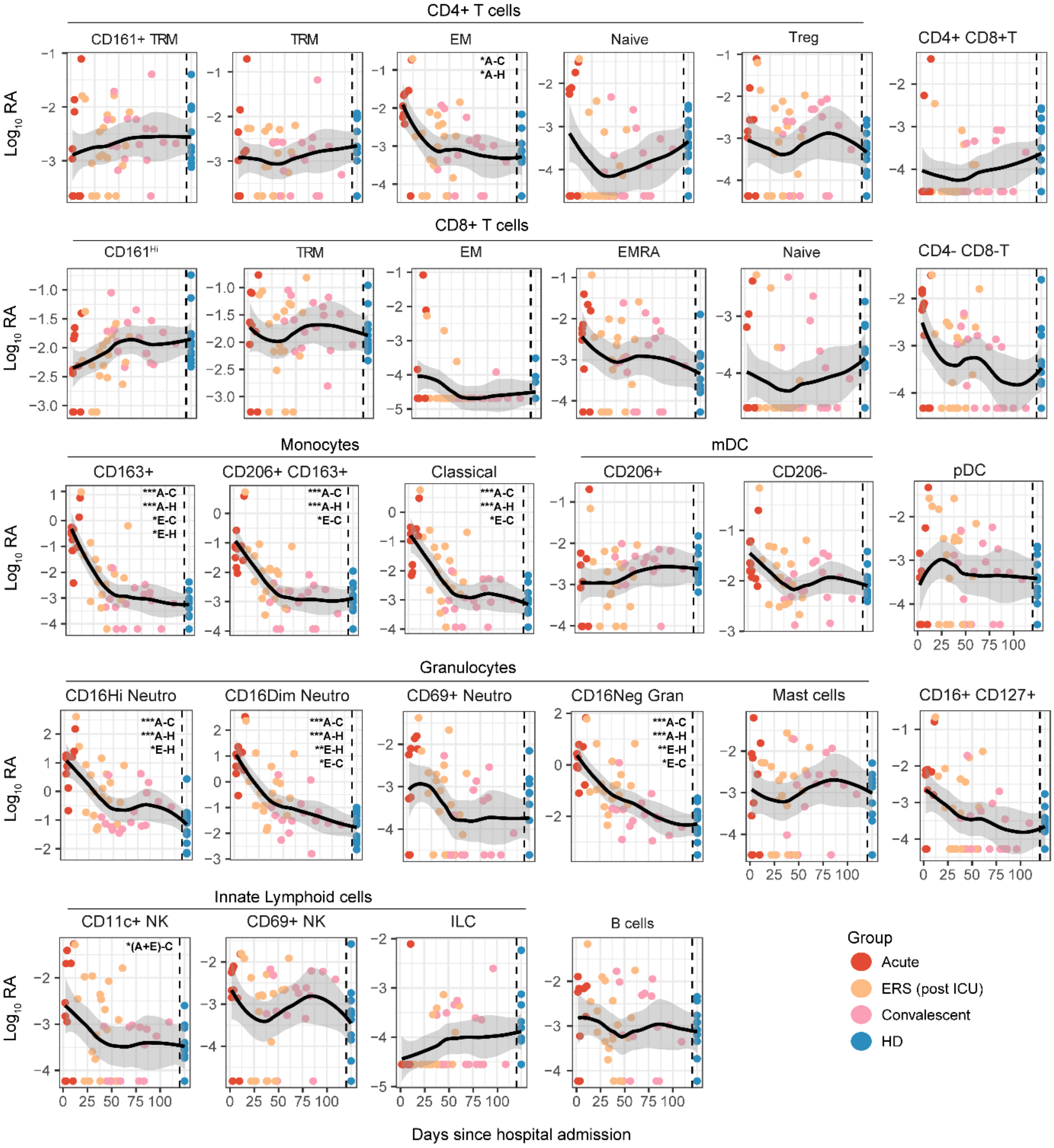


***Supplementary figure 1. Time plots of cell subsets relative to hospital admission.*** *Cell subset abundance during acute COVID-19 (red, 10 samples from 9 patients), during the early recovery phase (ERS, post ICU but still in hospital, orange, 18 samples from 11 patients n = 11), or in COVID-19 convalescence (5-6 weeks post hospital discharge, pink, n=16) or healthy controls (blue, n=12). Samples are plotted against day of hospital admission, with healthy donors plotted at the right axis separated by a dashed line. *p<0.05, **p<0.01, ***p<0.001, by linear-mixed model with group as fixed effect and individuals as random effect, with post-hoc testing and Tukey multiple testing correction followed by Benjamini-Hochberg correction for comparing multiple subsets. A = acute (hospitalized), E = early recovery stage, C = convalescent, 5-6 weeks post discharge, H = healthy donor. NK = natural killer cells. ILC = innate lymphoid cells, mDC = myeloid dendritic cells, pDC = plasmacytoid dendritic cells, Neutro = neutrophils, Trm = Tissue-resident memory, EM = effector memory, EMRA = effector memory re-expressing CD45RA. Treg = regulatory T cells.*

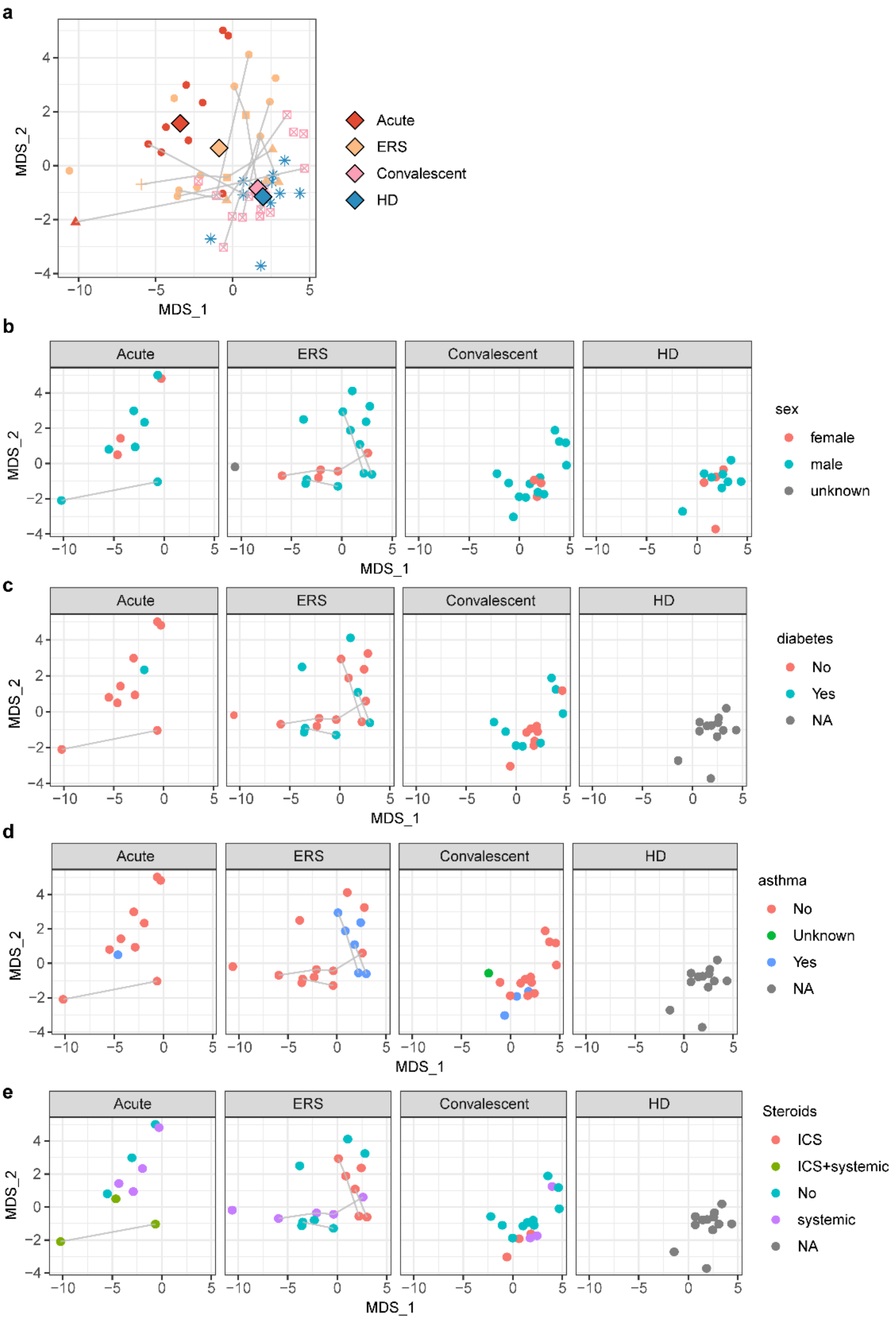


***Supplementary figure 2. Multi-dimensional scaling of all nasal subsets. a)*** *Clustering of all nasal samples based on abundance of all cellular subsets, colored by group. Acute COVID-19 (red, n=9), during the early recovery phase (ERS, post ICU, orange, n = 11), or in COVID-19 convalescence (5-6 weeks post hospital discharge, pink, n=16) or healthy controls (blue, n=12). Individual samples are shown with repeated samples per donor connected by lines. Centroids are shown per group as large diamonds. Per group faceted representation of the same scaling colored by covariates* ***b)*** *sex,* ***c)*** *diabetes,* ***d)*** *asthma,* ***e)*** *steroid usage. For these only repeated sample per group are shown and not across groups, e.g. from ERS to convalescence.*

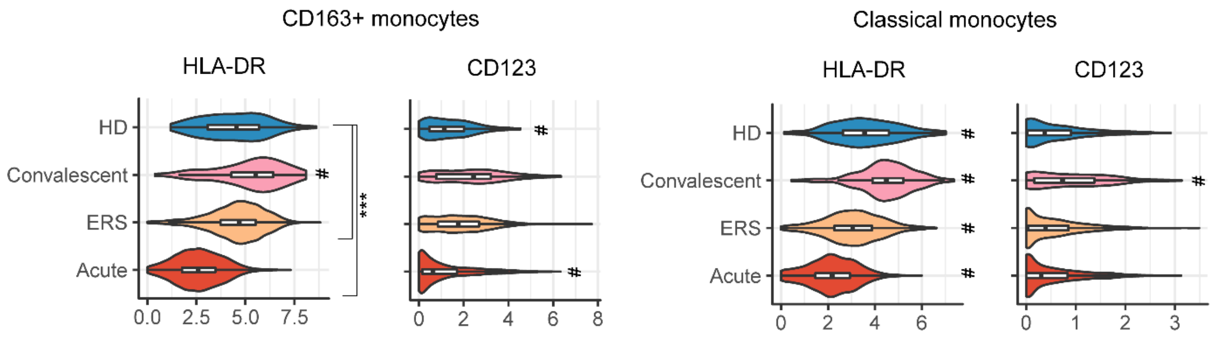


***Supplementary figure 3. Expression of HLA-DR and CD123 on CD163+ and CD163- monocytes.*** *Markers are shown as violin plots, with box plots inlayed per group. ***p<0.001, #p<0.001 compared to all other groups, using pairwise Wilcoxon test and Bonferroni correction for multiple testing. HD = healthy donors. ERS = early recovery stage.*

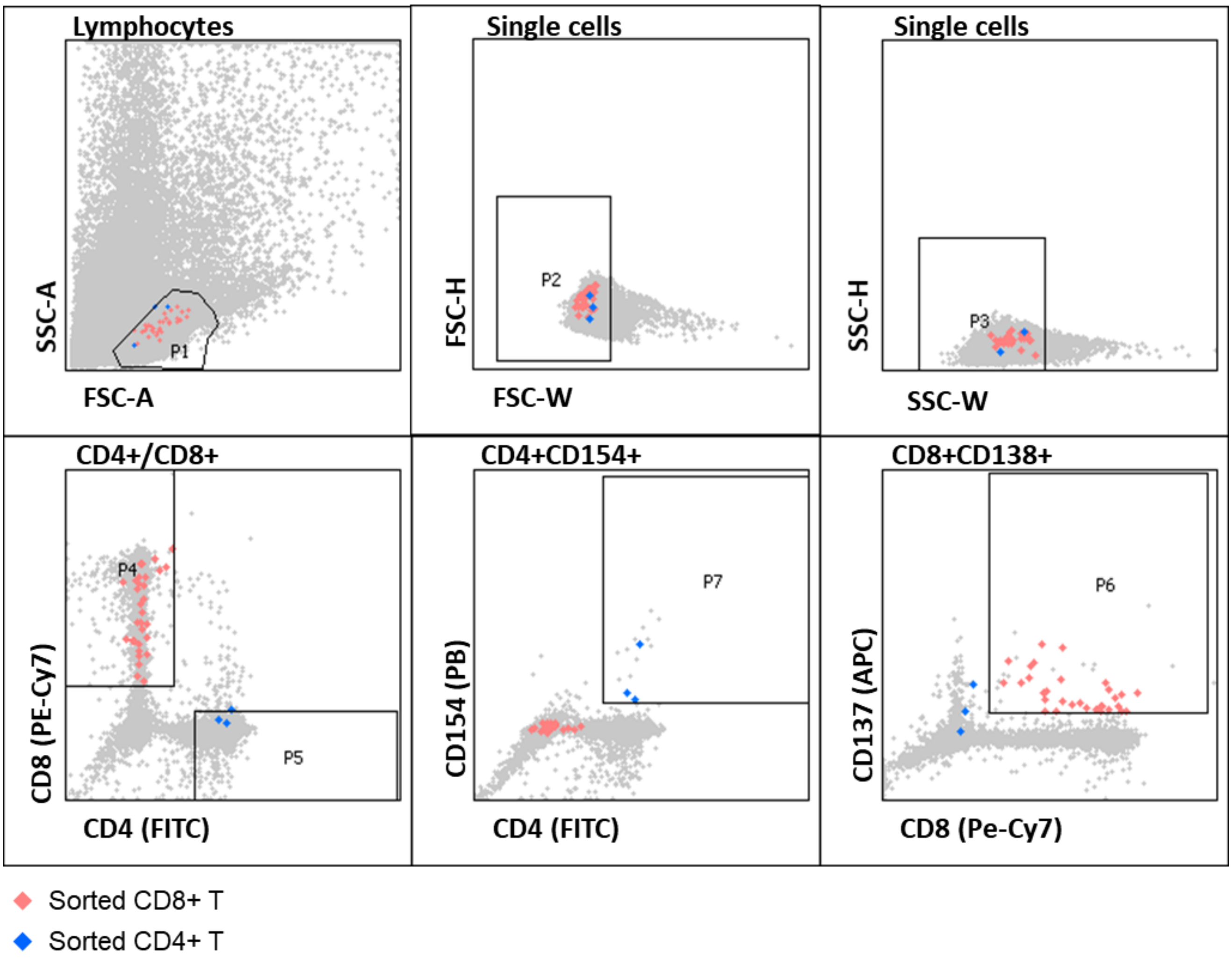


**Supplementary Figure 4. Sorting of SARS-CoV-2 specific T cells gating strategy.**

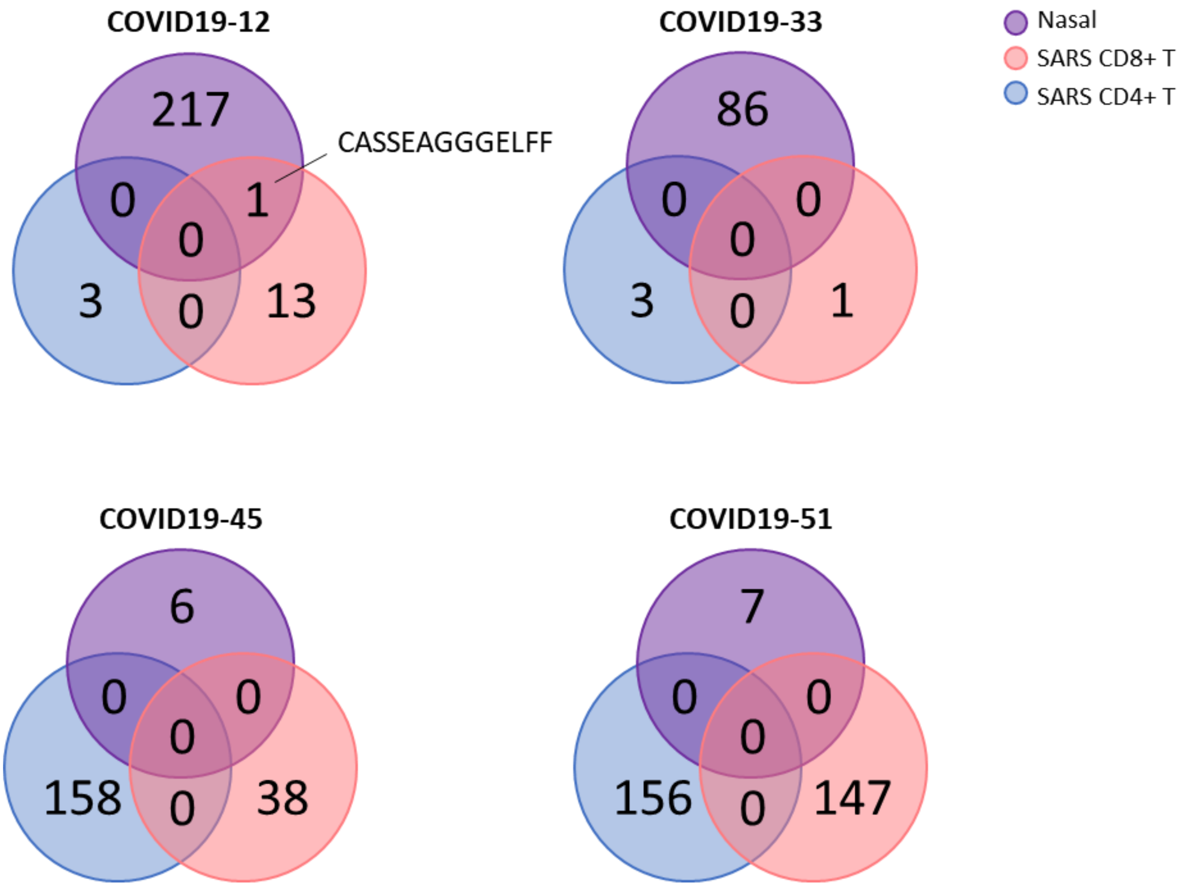


***Supplementary Figure 5. TCR clonotype numbers identified in nasal cells and sorted SARS-CoV-2 specific peripheral blood cells.*** *Each of the Venn diagrams represents the samples from a convalescent patient. The number of clones per cell type (total nasal cells, purple), SARS-CoV-2 specific CD8$^+$ T cells (red) and SARS-CoV-2 specific CD4$^+$ T cells (blue) are depicted in Venn diagrams for 4 individuals. An overlapping TCR was observed for patient 12 (top left), but not for the other 3 COVID-19 patients for which we attempted to match TCR sequences. This was likely due to the small TCR repertoire that was obtained in either the nasal scrapes (<10 clones for 2 individuals, bottom row) or SARS-CoV-2 reactive T cell subsets (<5 clones for 1 individual, top right).*

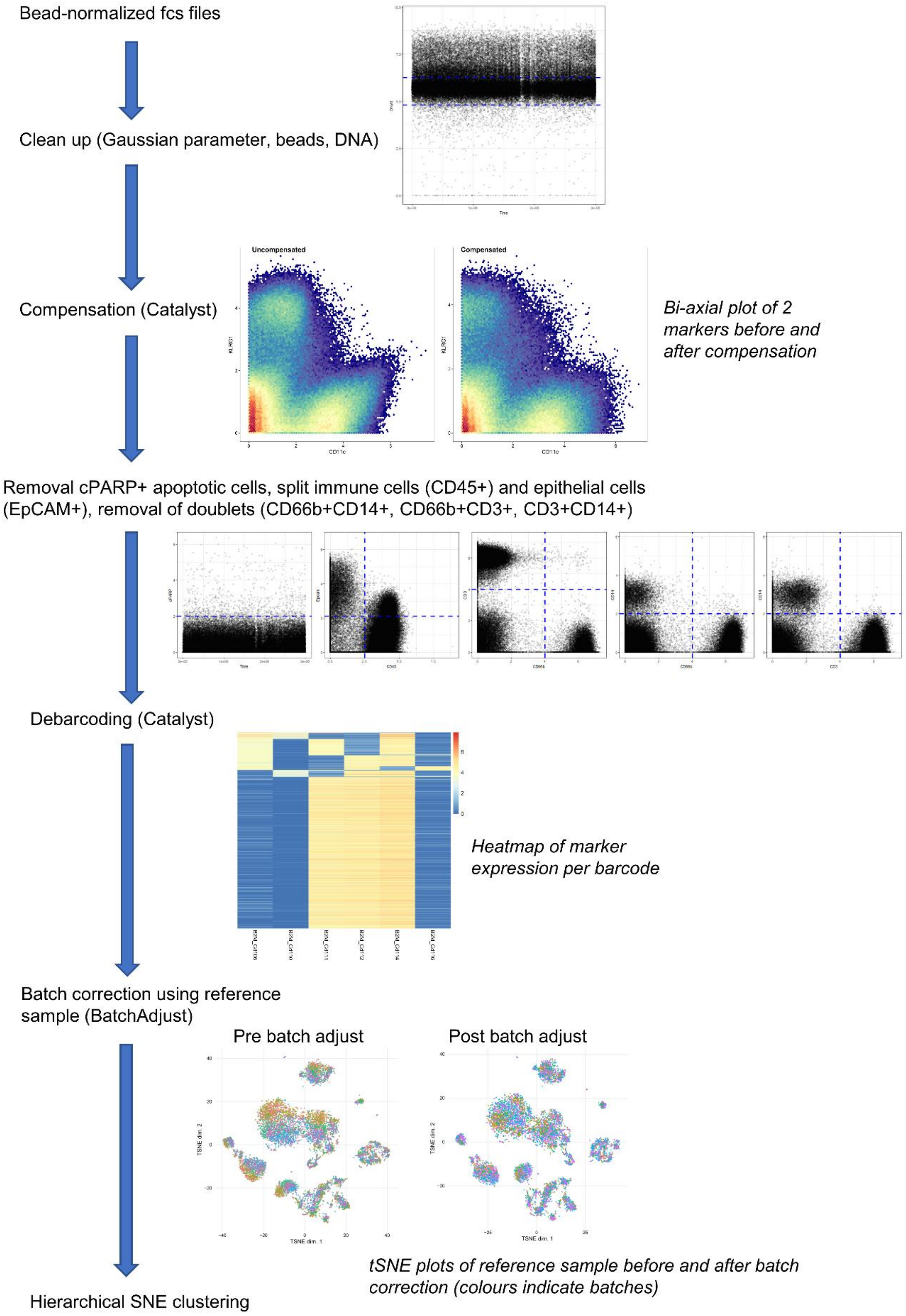


***Supplementary figure 6. CyTOF pre-processing pipeline.*** *Sequential steps in prior to data clustering are shown.*
